# Assessing the Pandemic Potential of Emerging Avian Influenza A(H5N1) in the United States Using the Viral Trait Assessment for Pandemics (ViTAP) Model

**DOI:** 10.1101/2025.02.04.25321643

**Authors:** Charles H. Jones, Marie Beitelshees

**Affiliations:** Independent Researcher, Tampa, FL 33602 USA; Bulmore Consulting, Lockport, NY 14094 USA

## Abstract

In an increasingly interconnected world, the threat of a devastating influenza pandemic looms large. Highly pathogenic avian influenza A(H5N1), with its high case fatality rate and potential for adaptation in avian reservoirs, is a prime candidate for triggering such a pandemic. We applied the Viral Trait Assessment for Pandemics (ViTAP) model, a quantitative framework, to assess the pandemic potential of H5N1 strains currently circulating in the United States. Through a systematic literature review and structured expert elicitation involving virologists and epidemiologists, we refined and validated ViTAP scores across 11 key viral traits. The resulting assessment yielded a final ViTAP score of 3.28 out of 5, placing the U.S. H5N1 strains in the moderate-to-high risk category. This elevated risk is driven by the virus’s high human fatality rate, its segmented RNA genome enabling reassortment, and its propensity for mutation. Sensitivity analyses highlight the transmission mode of the virus as a critical factor, with even minor increases in human-to-human spread dramatically escalating the pandemic threat. Comparisons with historical pandemics underscore the potential for H5N1 to rival the devastating impact of the 1918 H1N1 pandemic. Our findings emphasize the urgent need for enhanced surveillance in avian and mammalian hosts, targeted vaccine development, and comprehensive pandemic preparedness efforts. By demonstrating the utility of the ViTAP model for pandemic risk assessment, we provide a valuable tool to guide policy and interventions in the face of emerging zoonotic threats.

## Introduction

Influenza pandemics rank among the most devastating infectious disease events in human history, with the “Spanish flu” of 1918-1919 alone estimated to have claimed 50 million lives worldwide.^1^ The subsequent pandemics of the 20th century, while less severe, still exacted a heavy toll, causing an estimated 1-4 million deaths each.^2^ In the 21st century, the 2009 H1N1 “swine flu” pandemic demonstrated the speed with which a novel influenza virus can spread in an interconnected world, even if its virulence turned out to be comparatively mild.^3^

The pandemic threat posed by influenza A viruses stems from their high mutability and their segmented RNA genomes, which facilitate the rapid evolution of novel strains through mutation and reassortment.^4^ Aquatic birds are the natural reservoir for influenza A viruses, harboring a vast diversity of subtypes from which pandemic strains can emerge (**Figure *1*A**).^5^ Zoonotic transmission of avian influenza viruses from these wild birds to domesticated birds and other livestock is a key first step in this process, providing opportunities for the virus to spread to and adapt to human hosts and acquire the ability to spread efficiently from person to person. This is particularly troubling as H5N1 is currently spreading through domestic flocks in all 50 U.S. states with approximately 1,400 flocks impacted as of January 2025 (**Figure *1*B**).^6^

**Figure 1.**
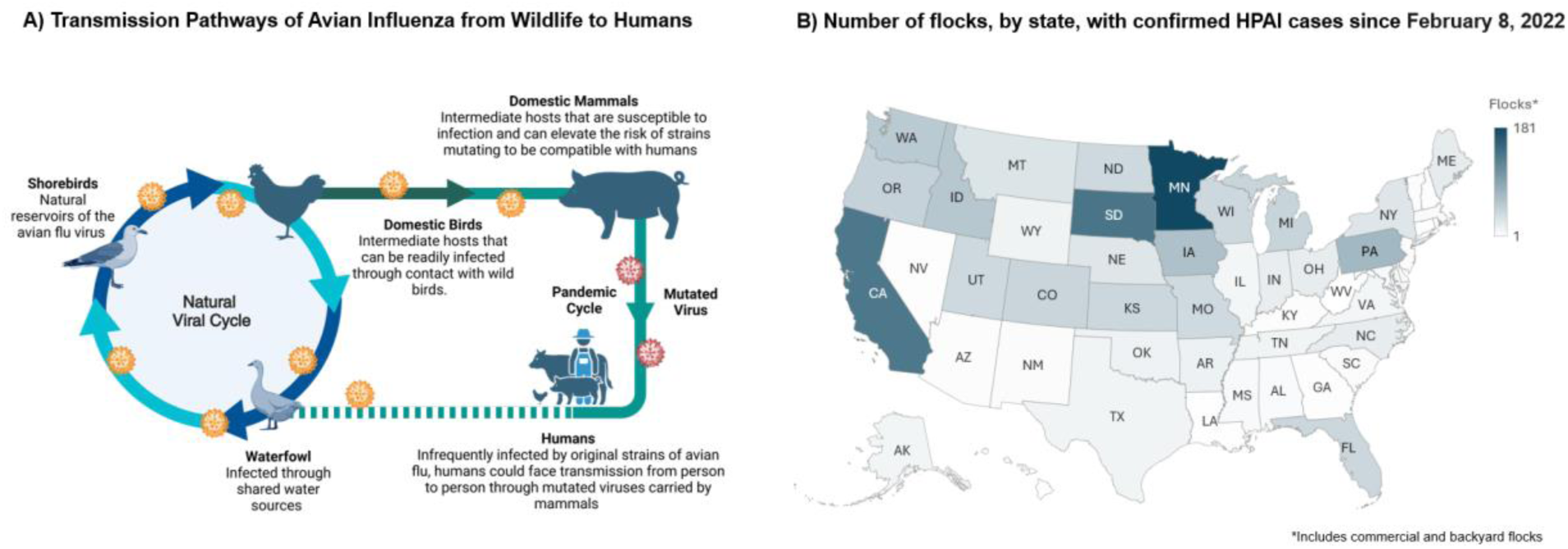
Transmission pathways and epidemiological spread among poultry of avian influenza in the United States. **A)** The pathways through which Avian Influenza transmits from wild birds, through domestic intermediates, to humans, highlighting the role of environmental and host factors in the evolution of pandemic-capable viral strains. Created in https://BioRender.com. State-specific counts of flocks with confirmed cases of highly pathogenic avian influenza (HPAI) as of February 8, 2022, showing the spatial distribution of outbreaks in the U.S., including commercial and backyard flocks. Data have been retrieved from USDA: Confirmations of Highly Pathogenic Avian Influenza in Commercial and Backyard Flocks | Animal and Plant Health Inspection Service (usda.gov)

Among avian influenza subtypes, A(H5N1) has been a top concern for pandemic preparedness since it first caused human infections in Hong Kong in 1997.^7^ The virus has since become enzootic in poultry across multiple countries in Asia and Africa, leading to sporadic but often severe human infections. From January 2003 to December 12, 2024, the World Health Organization (WHO) reported 954 confirmed human cases of H5N1, resulting in 464 deaths.^8^ This corresponds to a case fatality rate of approximately 49%, far exceeding that of seasonal influenza viruses (<0.1%)^9^ and even surpassing the 1918 pandemic virus (2-3%).^1^ Most recently, in January 2025, a death attributed to H5N1 was confirmed in Louisiana, marking the first H5N1-related human death in the United States.^10^

However, the true extent of human H5N1 infections may be greater than these figures suggest. Seroprevalence studies indicate that subclinical or mild H5N1 infections occur more frequently than previously recognized, particularly among populations with high exposure to poultry. A meta-analysis of H5 seroprevalence surveys in China estimated a seroprevalence of up to 3.2% in poultry workers.^11^ Similarly, a recent Centers for Disease Control and Prevention (CDC) study found detectable H5N1 antibodies in 7% of US dairy workers exposed to infected cattle, with half of the seropositive individuals reporting no symptoms.^12^ These findings suggest that the actual H5N1 infection fatality rate, while still high, may be lower than the observed case fatality rate.

Despite this broader spectrum of disease, the risk of H5N1 sparking a pandemic remains a serious concern. The segmented genome of H5N1 allows it to reassort with human influenza viruses, potentially acquiring gene segments that enhance transmissibility or alter antigenicity.^13^ Furthermore, the virus’s high mutation rate generates a diverse viral population that can rapidly adapt to new host environments.^14^ Several experimental studies have identified H5N1 strains with mutations that enable transmission via respiratory droplets or aerosols between ferrets, the gold-standard animal model for influenza transmissibility.^15,16^

To assess the pandemic risk posed by zoonotic influenza viruses like H5N1, the CDC developed the Influenza Risk Assessment Tool (IRAT),^17^ while the WHO created the Tool for Influenza Pandemic Risk Assessment (TIPRA).^18^ However, these tools have limitations: they heavily rely on epidemiological and clinical data that may be scarce in the early stages of an outbreak, they do not provide clear quantitative risk estimates, and their assessments are not routinely updated as new evidence emerges.

In this context, the Viral Trait Assessment for Pandemics (ViTAP) framework was developed as a novel, complementary approach for assessing the pandemic potential of zoonotic viruses.^19^ ViTAP quantifies the likelihood of a virus causing a pandemic based on its intrinsic characteristics, such as genomic structure, receptor binding, transmission mode, immune evasion, and virulence. Unlike other tools, ViTAP deliberately excludes external factors such as pre-existing immunity in human populations, the presence of therapeutics or vaccines, and geographical or environmental conditions. By focusing on traits inherent to the virus itself, ViTAP enables risk assessments even when epidemiological or clinical data are limited. Critically, ViTAP is designed to be a dynamic framework, with scores updated as new virological and epidemiological evidence becomes available.

Here, we present the first comprehensive assessment of the pandemic potential of H5N1 avian influenza using the ViTAP framework. We conducted an extensive literature review to gather data on the virus’s key traits and assign initial ViTAP scores. These scores were then refined through structured elicitation of input from leading experts in influenza virology, epidemiology, and public health. We assessed the sensitivity of the final scores to different assumptions about the relative importance of each trait. Finally, we compare H5N1’s ViTAP score to those of previous pandemic influenza viruses to provide context for the current level of risk.

Our findings aim to inform global efforts to prepare for and mitigate the risk of an H5N1 pandemic. By identifying the key viral traits that drive H5N1’s pandemic potential, we pinpoint crucial knowledge gaps and surveillance priorities. Importantly, we demonstrate how a rigorous yet accessible tool like ViTAP can provide actionable insights for pandemic preparedness, even for a virus as complex and long-studied as H5N1. As the threat of pandemic influenza persists, integrating such structured assessments into decision-making will be critical for safeguarding human lives and livelihoods worldwide.

## Methods

### ViTAP Framework

The ViTAP framework was originally developed as a comprehensive tool for assessing the pandemic potential of high-risk viruses, evaluating 14 key viral traits spanning genomic structure, epidemiological and clinical factors, and ecological interactions.^19^ Each trait was scored on a scale from 1 (low risk) to 5 (high risk), with the aggregate score calculated by weighting each trait according to its relative importance to pandemic risk. To compute an aggregate score for a given virus, each trait is assigned a weight reflecting its relative contribution to pandemic risk. The final ViTAP score is the sum of the products of each trait’s score and weight, yielding a number between 1 (lowest risk) and 5 (highest risk). Scores are interpreted as follows: 1-2 (low risk), 2-3 (medium risk), 3-4 (high risk), and 4-5 (very high risk). The full scoring rubric and weighting scheme are provided in the **Supplementary Materials**.

Importantly, ViTAP relies solely on intrinsic viral characteristics to calculate risk scores. External factors, such as population-level immunity, the availability of vaccines or antivirals, and socio-environmental conditions, are intentionally excluded from this framework. This ensures that ViTAP assessments remain consistent and focused on the biological properties of the virus itself. However, these scores can be contextualized by incorporating external data to provide a broader risk assessment and inform public health responses.

To adapt ViTAP for the assessment of H5N1, we streamlined the virus-agonistic framework (original ViTAP model) to focus on 11 traits with direct relevance to H5N1’s unique characteristics. Complex proxies for the virus’s ability to mutate and adapt, such as “Replication Machinery” and “Expected Impact of Mutations,” were consolidated into a single “Mutation Rate” category, as H5N1’s mutation rate is well-documented and measurable.^20–22^ Similarly, the “Segmented Genome” trait was integrated into the broader “Genome Type” category, reflecting the known significance of H5N1’s reassortment potential.^23–25^ As with the first generation ViTAP framework, a score between 1 (low risk) and 5 (high risk) is assigned for each trait based on predefined criteria derived from an extensive review of the virological and epidemiological literature.

This refinement process began with a comprehensive literature review to assign initial scores to each trait. We then engaged in a structured expert elicitation, convening a multidisciplinary panel to review the evidence, discuss discrepancies, and reach consensus on the final scores. Sensitivity analyses were conducted to evaluate the robustness of the scores under different weighting schemes and to identify the traits most influential in H5N1’s risk assessment.

These methodological adaptations demonstrate the flexibility of the ViTAP framework. By focusing on the most relevant traits for a given pathogen, the tool can be customized for targeted risk assessments while retaining its foundational principles. This iterative approach, combining literature review, expert input, and sensitivity analysis, ensures that ViTAP remains a dynamic and responsive tool for pandemic preparedness in the face of evolving viral threats.

### Literature Review and Initial Scoring

To inform the initial ViTAP scoring for H5N1, we conducted a systematic literature review to gather data on each of the 11 traits. We searched PubMed, Web of Science, and Google Scholar for studies published between 1997 (the year of the first recognized human H5N1 cases) and 2024. Search terms included “H5N1”, “avian influenza”, “highly pathogenic avian influenza”, “zoonotic influenza”, “pandemic risk”, “transmission”, “virulence”, “seroprevalence”, and related keywords.

We screened titles and abstracts to identify studies reporting primary data on H5N1’s epidemiological, clinical, and virological characteristics. We also included select review articles and meta-analyses that provided comprehensive summaries of the state of knowledge on specific traits. In total, we reviewed over 200 peer-reviewed publications, as well as reports from the WHO, CDC, and other public health agencies.

For each trait, we extracted key findings and used the predefined ViTAP scoring criteria to assign an initial score. When data were limited or ambiguous, we erred on the side of caution and assigned a higher score to reflect the inherent uncertainty.

### Expert Elicitation and Score Refinement

To ensure that the ViTAP scores for H5N1 reflected the most up-to-date and comprehensive understanding of the virus’s traits, we engaged in a structured expert elicitation process. We convened a panel of five experts in influenza virology, epidemiology, veterinary health, and pandemic preparedness. Participants were selected based on their publications, research funding, and contributions to national and international influenza initiatives.

The elicitation process involved three stages. First, each expert was provided with a summary of the ViTAP framework, the initial scores and justifications, and a subset of the key literature. Experts were asked to review these materials and provide written feedback on the appropriateness of each score, citing additional evidence as needed.

Next, we convened a series of videoconferences in which experts discussed their assessments and debated discrepancies. These discussions allowed for a nuanced consideration of the strengths and limitations of the available evidence, as well as an exploration of areas of uncertainty. Experts were encouraged to propose modifications to the scores or the scoring criteria in light of evolving virological and epidemiological insights.

Finally, experts completed a survey in which they provided their final scores and recommendations. The median of these individual scores was taken as the refined score for each trait. We assessed inter-rater reliability using Krippendorff’s alpha and addressed any residual discrepancies through follow-up discussions.

### Sensitivity Analysis

To assess the robustness of the final H5N1 ViTAP score, we conducted a sensitivity analysis in which we varied the weights assigned to each trait. We tested alternative weighting schemes that emphasized different categories of traits (e.g., genomic factors, transmissibility, virulence) and evaluated how these changes affected the overall score. We also performed a leave-one-out cross-validation, iteratively removing each trait from the model to assess its individual contribution to the final score.

In our comparison to prior pandemic viruses to contextualize H5N1’s pandemic potential, we used historical data to estimate ViTAP scores for the viruses responsible for the past four influenza pandemics: 1918 H1N1, 1957 H2N2, 1968 H3N2, and 2009 H1N1.^1–3^ For each virus, we reviewed the key literature on its origins, transmission patterns, clinical severity, and virological characteristics. While the available evidence is most limited for the oldest pandemics, this analysis provides a crude but informative benchmark for interpreting the scale of the risk implied by H5N1’s ViTAP score.

## Results and Discussion

### H5N1 ViTAP Scores and Evidence Summary

Our comprehensive analysis of H5N1’s pandemic potential reveals a concerning combination of viral traits that together indicate significant risk. Through systematic literature review and structured expert elicitation, we arrived at an overall ViTAP score of 3.28 out of 5 (**Table 1**), placing H5N1 firmly in the moderate-to-high risk category for pandemic potential. A full justification for individual characteristic scores is presented in the **Supplementary Information**.

**Table 1.** Final ViTAP trait scores for H5N1, incorporating both initial literature-based assessments and expert refinements.

| Category | Initial Score | Refined Score | Weight | Weighted Contribution |
| --- | --- | --- | --- | --- |
| Mutation Rate | 4.5 | 4.5 | 0.14 | 0.63 |
| Transmission Mode | 4.0 | 3.5 | 0.11 | 0.385 |
| Aerosol Transmission | 2.5 | 2.5 | 0.11 | 0.275 |
| Asymptomatic Spread | 1.5 | 1.5 | 0.11 | 0.165 |
| Human-to-Human Spread | 2.0 | 2.0 | 0.11 | 0.22 |
| Incubation Period | 4.0 | 4.0 | 0.05 | 0.20 |
| Symptom Severity | 5.0 | 5.0 | 0.06 | 0.30 |
| Mortality Rate | 5.0 | 5.0 | 0.06 | 0.30 |
| Demographic Impact | 3.0 | 3.0 | 0.04 | 0.12 |
| Genome Type | 5.0 | 5.0 | 0.12 | 0.60 |
| Reassortment Evidence | 4.0 | 4.0 | 0.02 | 0.08 |
| **Total** | – | – | – | **3.28** |

Several traits emerged as key drivers of H5N1’s elevated risk assessment, with its genomic characteristics being particularly significant. The virus received the maximum score of 5.0 for Genome Type, driven by its segmented RNA structure consisting of eight separate segments – a hallmark of all influenza A viruses.^26^ This genomic architecture enables reassortment events, where gene segments can be exchanged between different influenza strains during co-infection. The virus’s documented history of such events, both among avian strains and with human seasonal influenza viruses, resulted in a Reassortment Evidence score of 4.0.^23–25^ The pandemic risk posed by this reassortment capability is not theoretical – both the 1957 and 1968 influenza pandemics emerged through this process,^27^ demonstrating how quickly novel strains with pandemic potential can arise through genetic reassortment.

H5N1 scored high on measures of its evolutionary potential, with a Mutation Rate score of 4.5. Experts noted that this trait is key to the virus’s ability to continually escape population immunity and potentially adapt to sustained transmission in humans. Like other RNA viruses, H5N1 lacks proofreading mechanisms in its RNA polymerase, resulting in a range of mutations rates between 2.6 × 10^-3^ to 8.87×10^−3^ substitutions per site per year, dependent on host and strain.^20–22^ This range of rates enables rapid evolution and adaptation to new hosts and immune pressures, with billions of viral copies in a single infected host providing ample opportunity for beneficial mutations to arise.

The traits currently limiting H5N1’s pandemic potential are those associated with human transmission pathways. While the virus has demonstrated a well-documented ability to cross the species barrier from birds to humans, captured in its Transmission Mode score of 3.5 and evidenced by 954 confirmed human cases through December 2024,^8^ the consensus among experts was that H5N1 has not yet acquired the capacity for sustained person-to-person spread. This is reflected in the relatively low scores for both Aerosol Transmission (2.5) and Human-to-Human Spread (2.0). Though occasional instances of limited human-to-human transmission have been reported,^28^ the virus has not yet acquired the capacity for sustained transmission between people. However, several factors suggest this limitation could be overcome. First, the virus’s recent detection in mammalian hosts, including cattle,^12^ demonstrates its ongoing capacity to adapt to new species. Second, experimental studies using ferrets, the gold-standard animal model for human transmission, have shown that only a small number of mutations are required to enable respiratory droplet transmission.^15,16^ Experts agreed that these findings, combined with the increasing frequency of zoonotic spillover events, provide multiple opportunities for the virus to adapt to mammalian biology and potentially acquire more efficient human transmission capabilities.

The clinical impact of H5N1 infections also indicates a strong pandemic risk. The Mortality Rate received a score of 5.0, driven by the virus’s approximately 49% case fatality rate in confirmed human cases, which is orders of magnitude greater than that of seasonal influenza viruses.^8^ Even with the possibility of a substantial number of undetected mild infections,^11,12^ the consensus among experts was that H5N1’s fatality rate would still far exceed the pandemic benchmarks. Similarly, the Symptom Severity score of 5.0 underscores the virus’s capacity to cause severe disease. The ability to cause undetected infections is a key factor in a virus’s population spread, and there is currently limited evidence of asymptomatic spread of H5N1, resulting in a score of 1.5. However, based on growing seroprevalence data,^11,12^ experts cautioned that H5N1’s high seroprevalence in some exposed groups means that its true incidence may be considerably higher than the reported case counts suggest.

The Incubation Period received a score of 4.0, based on estimates of 2 to 5 days in confirmed cases.^29^ This duration is longer than that of seasonal influenza but shorter than some other zoonotic viruses, providing enough time for potential spread before symptom onset while still allowing for effective contact tracing and containment measures.

The Demographic Impact score of 3.0 reflects the virus’s disproportionate impact on children and young adults, with a median age of 18 years.^28^ This pattern is particularly concerning, as high morbidity and mortality in younger age groups can have severe societal and economic consequences. However, the overall demographic impact remains limited by the small total number of cases. Should the virus acquire efficient human-to-human transmission capability, this impact could increase significantly.

Taken together, these trait scores paint a troubling picture of H5N1’s pandemic potential. On one hand, the virus’s current lack of efficient human-to-human transmission is a limiting factor. But its high mutability, frequently reassorting genome, and history of steady spillovers into humans all provide a rich set of evolutionary pathways by which human transmissibility could emerge. This potential is made all the more concerning by the virus’s severity; even a partially-attenuated H5N1 strain that retained a fraction of the observed case fatality rate could have a devasting global impact if it spread efficiently between people.

### Sensitivity Analysis of H5N1 Scores

The sensitivity analysis (**Table 2**) reveals a clear hierarchy in how different traits influence H5N1’s overall pandemic potential. By systematically varying the weights of individual traits by ±20%, we identified which characteristics most strongly influence the final ViTAP score and, by extension, our assessment of pandemic risk.

**Table 2.**
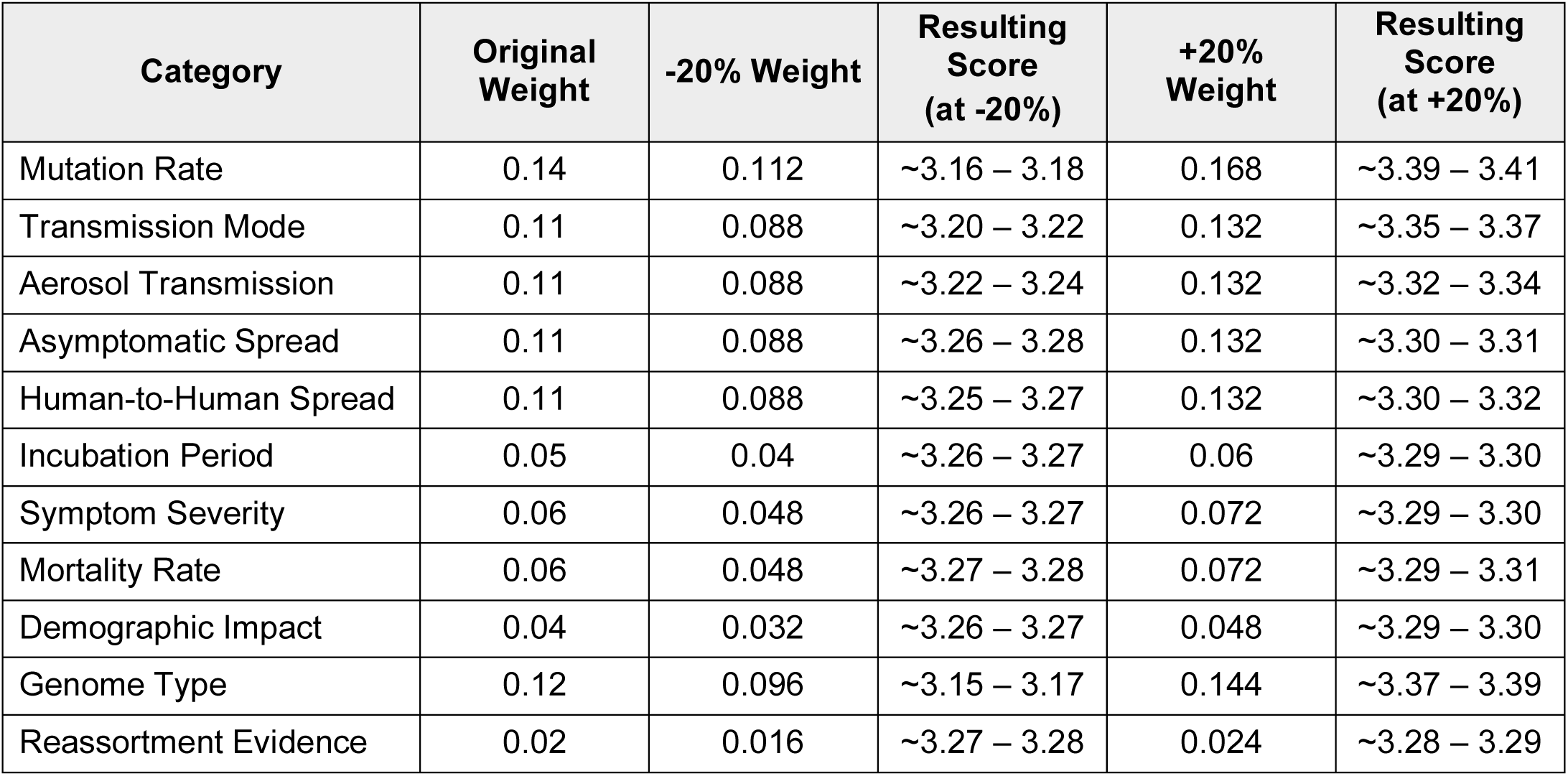
Sensitivity analysis results showing the impact of varying trait weights on the final H5N1 ViTAP score.

| Category | Original Weight | -20% Weight | Resulting Score (at -20%) | +20% Weight | Resulting Score (at +20%) |
| --- | --- | --- | --- | --- | --- |
| Mutation Rate | 0.14 | 0.112 | ~3.16 – 3.18 | 0.168 | ~3.39 – 3.41 |
| Transmission Mode | 0.11 | 0.088 | ~3.20 – 3.22 | 0.132 | ~3.35 – 3.37 |
| Aerosol Transmission | 0.11 | 0.088 | ~3.22 – 3.24 | 0.132 | ~3.32 – 3.34 |
| Asymptomatic Spread | 0.11 | 0.088 | ~3.26 – 3.28 | 0.132 | ~3.30 – 3.31 |
| Human-to-Human Spread | 0.11 | 0.088 | ~3.25 – 3.27 | 0.132 | ~3.30 – 3.32 |
| Incubation Period | 0.05 | 0.04 | ~3.26 – 3.27 | 0.06 | ~3.29 – 3.30 |
| Symptom Severity | 0.06 | 0.048 | ~3.26 – 3.27 | 0.072 | ~3.29 – 3.30 |
| Mortality Rate | 0.06 | 0.048 | ~3.27 – 3.28 | 0.072 | ~3.29 – 3.31 |
| Demographic Impact | 0.04 | 0.032 | ~3.26 – 3.27 | 0.048 | ~3.29 – 3.30 |
| Genome Type | 0.12 | 0.096 | ~3.15 – 3.17 | 0.144 | ~3.37 – 3.39 |
| Reassortment Evidence | 0.02 | 0.016 | ~3.27 – 3.28 | 0.024 | ~3.28 – 3.29 |

Three traits emerged as primary drivers of score variation. Mutation Rate demonstrated the largest impact, with score fluctuations between 3.16 and 3.41 (Δ = 0.25) when adjusting its weight. This pronounced influence corresponds to the fundamental role of mutation in viral adaptation to new hosts and escape from population immunity. Genome Type showed similarly substantial impact (range: 3.15 - 3.39, Δ = 0.24), a finding consistent with how the segmented nature of the influenza genome facilitates rapid evolution through reassortment. Transmission Mode, which for H5N1 is currently zoonotic transmission, exhibited the third-highest impact (range: 3.20 - 3.37, Δ = 0.17). This substantial variation is particularly concerning given H5N1’s current transmission score of 3.5 primarily represents zoonotic spread. Should the virus acquire mutations enabling efficient human-to-human transmission via respiratory droplets – the typical route for human influenza viruses - the transmission score would increase to 5.0, potentially shifting the overall ViTAP score well into the high-risk category.

We observed moderate influence for Aerosol Transmission (Δ = 0.12) and Human-to-Human Spread (Δ = 0.07). The interplay between these traits and Transmission Mode is critical – while current human transmission remains limited, any viral adaptations that enhance airborne aerosol spread between humans could dramatically increase pandemic risk.

Several traits traditionally associated with disease severity showed surprisingly minimal impact on the overall score. Mortality Rate, Symptom Severity, and Demographic Impact all produced score variations of 0.04 or less when their weights were adjusted. This result suggests that while these factors are crucial for public health impact and clinical outcomes, they may be less critical in the case of H5N1 than evolutionary and transmission characteristics in determining pandemic potential.

Reassortment Evidence showed the smallest score variation (Δ = 0.02), though this requires careful interpretation within the context of the ViTAP framework. The minimal variation stems from the trait’s intentionally low weight (0.02) in the model, as it functions primarily as a modifier for the more heavily weighted Genome Type trait (0.12). This design choice acknowledges that while reassortment events can drive dramatic phenotypic changes, the capacity for reassortment is already partially captured in the Genome Type score for segmented RNA viruses like influenza A. The high scores for both Reassortment Evidence (4.0) and Genome Type (5.0) in H5N1 are supported by the virus’s documented history of reassortment and its inherent genomic capability for such events.

These findings have significant implications for surveillance and pandemic preparedness efforts. The analysis indicates that monitoring programs should prioritize tracking mutations and transmission patterns, as changes in these parameters could most dramatically affect H5N1’s pandemic potential. The substantial impact of Genome Type variations also establishes the importance of monitoring reassortment events, particularly in regions where multiple influenza strains co-circulate in avian or mammalian hosts.

It is important to note that this sensitivity analysis only examines the impact of changing the trait weights, not the scores themselves. The scores are based on the best available evidence and expert judgment and should be updated as new information becomes available. The goal of the sensitivity analysis is to provide insight into which traits have the greatest influence on the overall assessment, and to help identify areas where additional data could most improve the reliability of the ViTAP framework. Importantly, despite the significant weight variations tested in this analysis, the final ViTAP score remained within the moderate-to-high risk range (3.15 - 3.41). This stability across different weighting schemes supports the robustness of the framework’s risk categorization and increases confidence in the overall assessment of H5N1’s pandemic potential.

### Contextualization of H5N1 ViTAP Scores to Previous Influenza A Pandemics Strains

Comparisons with previous pandemic influenza viruses provide important context for interpreting H5N1’s ViTAP score. To accomplish this, the numerical scores across the 11 traits for each pathogen were taken from our initial study^19^ and run through the ViTAP framework with revised weightings (i.e., the model used to calculate the H5N1 score) to ensure consistency and accurate comparisons. The results of the analysis are presented in **Figure 2**.

**Figure 2.**
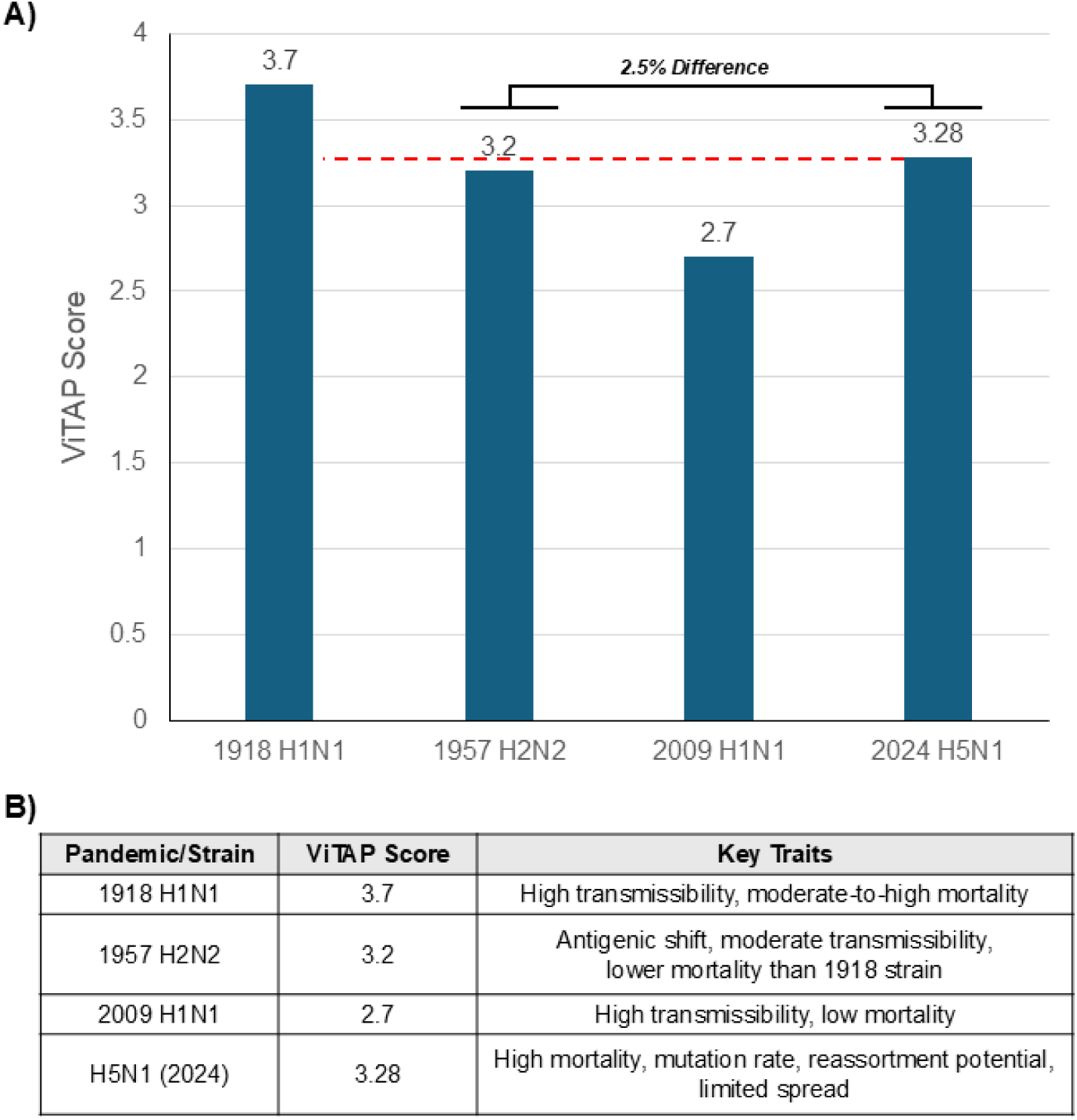
Approximate ViTAP Scores for Historical Influenza Pandemics. **A)** ViTAP scores calculated using revised weighting and valuation illustrate the pandemic potential of historical and emerging influenza strains. The dashed red line highlights the score differences across all included influenza pandemics. B) Approximate ViTAP scores and key traits for historical influenza pandemics demonstrate differences in pandemic potential and viral characteristics.

H5N1’s ViTAP score was found to be 3.28, 2.5% higher than the 1957 H2N2 influenza pandemic, which had a score of 3.2 and which caused an estimated 1-4 million deaths worldwide (**Figure 2A**).^2^ This increase reflects H5N1’s higher intrinsic pandemic potential based on its virological characteristics, including its segmented genome, high mutation rate, and potential for zoonotic transmission (Figure 2B).

We then applied the score adjustment to the virus-family-agnostic risk categorization system described in Jones et al. (2024), which was inspired by the hurricane risk classification system. Using this approach, the 2024 H5N1 strain’s adjusted score of 3.28 places it squarely in the highest risk category (Category 6), alongside the 1957 influenza pandemic, the COVID-19 pandemic (SARS-CoV-2), and the 1918 influenza pandemic (**Supplementary Figure 1**). This suggests that H5N1 has potential to cause a severe pandemic, potentially on the order of the catastrophic 1918 Flu, if it overcomes its current limitations in sustained human-to-human transmission.

By applying this process to any virus in a category beyond influenza, the ViTAP model could serve as a foundational tool for evaluating pandemic potential in other viral families. For example, by adapting the scoring criteria and weights to account for the unique genomic, transmission, and virulence traits of coronaviruses, paramyxoviruses, or filoviruses, the model can provide a standardized risk assessment framework. This approach ensures comparability across diverse pathogens, enabling the identification of high-risk viruses within and across families. Moreover, these virus-specific models can be benchmarked against historical pandemics, as demonstrated with H5N1, allowing their risk to be contextualized within a broader public health risk categorization system.

Of course, such comparisons have limitations. The ViTAP framework attempts to quantify pandemic risk based on current virological and epidemiological traits, but it cannot perfectly predict how these traits might change as a virus evolves. Nor can it capture the myriad ecological, demographic, and behavioral factors that also shape a pandemic’s course. Nonetheless, by focusing the attention of the scientific and public health community on the specific viral characteristics that make H5N1 such a legitimate threat, the ViTAP assessment can help guide priorities for prevention and response.

Foremost among these is the need for intensive, coordinated surveillance efforts to track the ongoing evolution of H5N1 across the human-animal interface. From an epidemiological perspective, this means enhancing systems for rapidly detecting human infections, including protocols for contact tracing and the examination of clusters to distinguish between human-to-human transmission and common source exposures. It also demands expanded surveillance and sampling in avian populations, wild and domestic, to generate high-resolution genomic and phenotypic data. Coupling this work with routine screening of other animals, like swine and cattle, that could serve as bridging hosts will be vital for detecting any crossovers that might presage a shift toward mammalian adaptation.

On the virological front, a key priority is to better characterize the specific mutations and reassortments that could render H5N1 more transmissible in humans. Gain-of-function experiments, conducted under appropriate biosafety controls, may be justified to assess the phenotypic impacts of key molecular changes. Complementary studies using *in vitro* and *in vivo* models can shed light on the host range, cell tropism, receptor binding, and replication efficiency of H5N1 variants. Integrating these experimental insights with computational models and machine learning techniques could help predict which constellations of viral traits are most likely to support a human-adapted strain.

To illustrate this concept, let us consider a hypothetical scenario. Imagine that through surveillance, a new H5N1 strain is detected in poultry that has acquired a mutation in its hemagglutinin (HA) gene known to enhance binding to human-type receptors. This finding would immediately raise concerns about the virus’s potential for mammalian adaptation. To assess this risk, researchers could conduct a series of experiments:

1. Receptor binding assays to quantify the mutant virus’s affinity for human vs. avian receptors
2. *In vitro* replication studies in human airway epithelial cells to assess the virus’s ability to infect and spread in human tissues
3. Ferret transmission studies to evaluate the virus’s ability to spread via respiratory droplets in a mammalian model
4. Computational modeling to predict the structural and functional consequences of the HA mutation and its potential to interact synergistically with other adaptive changes

By integrating data from these complementary approaches, researchers could develop a more comprehensive understanding of the functional significance of the HA mutation and its implications for the virus’s pandemic potential. This understanding, in turn, could inform decisions about vaccine strain selection, antiviral stockpiling, and public health interventions.

This is just one illustrative example of how a multidisciplinary research approach can help elucidate the complex interplay of factors that shape a virus’s pandemic risk. In reality, assessing the pandemic potential of a virus like H5N1 requires a sustained, collaborative effort that draws on insights from virology, epidemiology, immunology, veterinary medicine, and public health.

The development of effective medical countermeasures is another critical avenue for mitigating the risk posed by H5N1. While a number of H5N1 vaccine candidates have been developed and tested in clinical trials,^30^ the rapid evolution of the virus means that a closely matched vaccine may not be immediately available in the event of a pandemic. Efforts to improve the speed of vaccine development, including the use of novel platforms like mRNA vaccines, could help close this gap. Similarly, the identification and preclinical testing of antiviral drugs and monoclonal antibodies with activity against H5N1 may provide valuable tools for treatment and prophylaxis.

Beyond these biomedical interventions, it is crucial that we apply the lessons learned from the COVID-19 pandemic to strengthen the global public health infrastructure for responding to potential future threats like H5N1. This includes bolstering the capacity for rapid diagnostic testing, enhancing systems for the collection and sharing of epidemiological data, and investing in risk communication strategies to combat misinformation and promote effective public health measures. It also means addressing the profound global inequities in access to preventative and therapeutic interventions – a moral imperative that is also a fundamental matter of collective biosecurity.

Continued refinement of tools like ViTAP, and their integration into decision-making processes, can play a valuable role in focusing these efforts. As our understanding of the viral determinants of pandemic potential continues to grow, it will be important to update and validate the ViTAP scoring criteria. Expanding the framework to incorporate ecological and host factors, in addition to intrinsic viral traits, could provide an even more nuanced picture of the risk landscape. And as demonstrated by this H5N1 assessment, using ViTAP as a focal point for structured expert elicitation can be a powerful approach for building scientific consensus and identifying key knowledge gaps.

Moreover, the process used to refine the ViTAP model for influenza viruses – modifying categories and adjusting weights through systematic expert review – can be extended to develop similar models for other classes of pathogens, such as bacteria and fungi. For bacterial pathogens, a BacTAP (Bacterial Trait Assessment for Pandemics) model could be constructed by identifying relevant traits, such as antimicrobial resistance, environmental resilience, and virulence factors. Similarly, a FunTAP (Fungal Trait Assessment for Pandemics) model could incorporate traits like spore production, environmental adaptation, and immunoevasive capabilities. These models would require comprehensive literature reviews to define the traits most relevant to each pathogen class and their weights. Structured expert elicitation, involving microbiologists, infectious disease specialists, and epidemiologists, would then refine these weights and scoring criteria, ensuring accuracy and relevance. This iterative process would allow the development of pathogen-class-specific models that are aligned with ViTAP’s dynamic framework, enabling consistent and comparable risk assessments across diverse microbial threats.

Ultimately, the reality is that for all our scientific and technological advances, we remain profoundly vulnerable to pandemic threats like H5N1. Influenza viruses are masterful in their ability to evolve and adapt, and the increasing pressures of globalization, climate change, and habitat encroachment only serve to accelerate their opportunities to do so. In this context, complacency is not an option. Only by sustaining a posture of vigilance and continually working to improve our understanding of the viral factors that enable pandemics, can we hope to stay one step ahead of the next global health crisis. The assessment of H5N1 using the ViTAP framework is an important step in this direction – but it is only one small piece of the comprehensive, collaborative response that will be required to safeguard our collective future.

## Conclusion

This study presents the first comprehensive assessment of avian influenza A(H5N1)’s pandemic potential using the Viral Trait Assessment for Pandemics (ViTAP) framework. The final ViTAP score of 3.28, derived from extensive literature review and structured expert elicitation, places H5N1 in the moderate-to-high risk category. This elevated risk profile arises from the complex interplay of H5N1’s high human case fatality rate, segmented RNA genome prone to reassortment, high mutation rate, and propensity for zoonotic spillover. Sensitivity analysis highlights the critical finding that seemingly modest changes in transmission characteristics could rapidly escalate pandemic risk, even while other viral properties remain stable.

These results have significant implications for pandemic preparedness strategies. The traditional approach of monitoring individual viral characteristics must evolve into integrated surveillance systems capable of detecting and responding to concurrent changes across multiple traits. Comparisons with previous pandemic influenza viruses underscore the gravity of the H5N1 threat: while not yet at the level of the catastrophic 1918 H1N1 virus, H5N1’s ViTAP score aligns closely with the 1957 H2N2 virus, which claimed an estimated 1-4 million lives worldwide. In our globalized era of high human interconnectedness, an H5N1 virus that achieves even a fraction of its current case fatality rate with efficient human transmission could have devastating consequences.

Urgent action is needed to mitigate this risk. Key priorities include:

1. Expanding genomic and phenotypic surveillance in avian, swine, and human populations to detect mutations and reassortment events that could signal a shift towards mammalian adaptation and transmission.
2. Conducting experimental studies to elucidate the virological determinants of H5N1’s host range, receptor binding, transmissibility, and virulence, informing predictive models of the virus’s evolutionary trajectories.
3. Accelerating the development and testing of broadly protective, rapidly scalable H5N1 vaccines and therapeutics.
4. Strengthening global public health infrastructure for pandemic preparedness and response, emphasizing equitable access to countermeasures and proactive risk communication.

The path forward demands a fundamental shift in our conceptualization of and preparation for pandemic threats. Surveillance systems must incorporate real-time monitoring of viral genetic and phenotypic changes. The ViTAP framework, by providing standardized, quantitative risk assessment, can focus scientific attention and resources on the most critical aspects of viral evolution and adaptation. This systematic, evidence-based approach, grounded in expert elicitation from diverse disciplines, is crucial for protecting global public health against emerging pandemic threats. As H5N1 continues to evolve, such cross-disciplinary collaboration will be increasingly vital to keep pace with the virus’s epidemic and pandemic potential.

## Data Availability

All data produced in the present work are contained in the manuscript

## Author Contributions

CHJ conceptualized and designed the study, guided the execution of the project and contributed in equal parts to the writing. MB assisted with the execution of the study, provided strategic oversight and direction to the execution of the study and contributed in equal parts to the writing.

## Correspondence

Correspondence and requests for materials should be addressed to

## Supplementary Information

### 1. Extended Methods

#### 1.1. Expert Selection and Questionnaire Process

##### Expert Panel Composition

Five experts were invited to refine the initial H5N1 Viral Trait Assessment for Pandemics (ViTAP) scores – three virologists specializing in influenza virology and two epidemiologists with a focus on zoonotic diseases. Criteria for inclusion in this panel were:

1. At least five years of active research on avian influenza or related RNA viruses.
2. Experience with analyzing or modeling pandemic threats (e.g., influenza risk assessment, SARS-CoV-2 modeling).
3. Willingness to participate in a structured discussion to reconcile diverging trait scores.

##### Questionnaire Outline

Prior to the consultation, each expert received a structured document titled “**H5N1 ViTAP Questionnaire**,” containing:

- **Background on the ViTAP Model**: A concise explanation of the 11 ViTAP categories, scoring scales (1–5), and typical weighting scheme.
- **Preliminary H5N1 Scores**: For each category, a table showed the *initial* literature-based score, a short rationale, and key data references (e.g., WHO bulletins, peer-reviewed publications).
- **Discussion Points**: Targeted queries, such as:
- *“Does currently available data justify a Transmission Mode score >4.0, or is 3.5 more consistent with contact-driven spread?”*
- *“Should Mortality Rate be adjusted to account for possible underreporting of mild H5N1 cases?”*

The final page solicited *open-ended comments* on each category’s justification, with a request to highlight any major uncertainties or newly published evidence not captured in the preliminary scoring.

##### Refinement Protocol

Experts returned the questionnaire within one week, indicating for each category whether they agreed or disagreed with the preliminary score (±0.5). Where there was disagreement (≥2 experts diverging by >1 point), a 60-minute virtual panel discussion was scheduled. Scores were updated only after at least four experts arrived at consensus within ±0.5 of the final value.

#### 1.2. Weighted Scoring Overview

The ViTAP model uses a weighting scheme adapted from Jones et al. (PNAS Nexus 2024).^1^ Each trait’s relative weight is:

- **Mutation Rate**: 0.14
- **Transmission Mode**: 0.11
- **Aerosol Transmission**: 0.11
- **Asymptomatic Spread**: 0.11
- **Human-to-Human Spread**: 0.11
- **Incubation Period**: 0.05
- **Symptom Severity**: 0.06
- **Mortality Rate**: 0.06
- **Demographic Impact**: 0.04
- **Genome Type**: 0.12
- **Reassortment Evidence**: 0.02

In some contexts, weights are normalized to sum to 1.0 exactly. However, for the present H5N1 evaluation, they sum to ∼0.93. This minor discrepancy (0.93 vs. 1.0) arises from historical calibration across multiple viruses and does not significantly affect interpretability of the final composite. By design, categories like Genome Type (segmented vs. non-segmented) and Mutation Rate are heavily weighted for influenza-like viruses.

### 2. Justifications for Each Initial ViTAP Trait Score Assigned to H5N1

#### Genome Type (Initial Score: 5.0)

Influenza A viruses like H5N1 have a segmented RNA genome, which is the highest risk category for pandemic potential in the ViTAP framework. The segmented nature of the genome allows for reassortment events, where gene segments can be exchanged between different influenza strains during co-infection of a host cell. This process can lead to the rapid emergence of novel strains with pandemic potential, as seen in the 1957 and 1968 influenza pandemics.^2^

To understand why this is such a high-risk trait, let us consider an analogy. Imagine that a virus’s genome is like a recipe book, and each gene segment is like a page with instructions for making a specific dish. In a virus with a non-segmented genome, all the pages are bound together, so it is difficult to make changes to the recipe. But in a segmented genome, the pages are loosely bound, making it easier to swap out a page (i.e., a gene segment) from a different recipe book. This mixing and matching of pages can result in a new “cookbook” with recipes for dishes that have never been seen before - in virological terms, a novel strain with unknown properties.

#### Symptom Severity (Initial Score: 5.0)

In contrast to the uncertainty around asymptomatic infection, the severity of symptoms in confirmed H5N1 cases is well documented. The majority of patients develop severe respiratory disease, often progressing to acute respiratory distress syndrome and multi-organ failure.^3^ Neurological complications, including encephalitis, have also been reported.^3^ While it is possible that milder cases are being missed, the high proportion of severe outcomes among confirmed cases warrants a high score for this trait.

The severity of H5N1 infections is a major factor in its pandemic risk. A virus that causes severe disease can quickly overwhelm healthcare systems, leading to high morbidity and mortality. This is particularly true in resource-limited settings, where access to advanced medical care may be scarce. Even in well-resourced healthcare systems, a severe pandemic can strain capacity and lead to difficult triage decisions. The 1918 influenza pandemic, which had a case fatality rate of 2-3%, provides a sobering example of the potential impact of a severe pandemic virus.^4^

#### Mortality (Initial Score: 5.0)

H5N1 has an exceptionally high case fatality rate in humans, with 464 deaths reported among the 954 confirmed cases to date (49.4%).^5^ This is one of the highest mortality rates among known zoonotic pathogens and is orders of magnitude higher than typical seasonal influenza viruses. Even if the true mortality rate is lower due to undetected mild cases, it would still far exceed the pandemic severity thresholds used in the ViTAP framework. Therefore, we assigned the maximum score of 5 for this trait.

To put H5N1’s mortality rate in context, let us compare it to some other notable pathogens. The case fatality rate for SARS-CoV-2, the virus responsible for the COVID-19 pandemic, is estimated to be around 1-2%.^6^ For MERS-CoV, another zoonotic coronavirus, the case fatality rate is around 34%.^7^ The 1918 influenza pandemic, as mentioned earlier, had a case fatality rate of 2-3%.^4^ H5N1’s mortality rate of nearly 50% in confirmed cases is staggering in comparison. Even if this is an overestimate due to case ascertainment bias, it highlights the extraordinary virulence of this virus.

#### Mutation Rate (Initial Score: 4.5)

Like other RNA viruses, influenza A viruses have a high mutation rate due to the lack of proofreading mechanisms in their RNA polymerase.^8^ This allows for rapid evolution and adaptation to new hosts and immune pressures. While H5N1 does not have the highest mutation rate among influenza subtypes, its rate is still substantial and contributes to its pandemic potential. Studies have estimated the mutation rate of H5N1 to be between 2.6 × 10^-3^ to 8.87×10^−3^ substitutions per site per year.^9–11^

To put this in perspective, let us think about what a mutation rate of 2.6 × 10^-3^ substitutions per site per year means. In a virus with a genome of approximately 13,500 nucleotides (like H5N1), this rate would translate to around 35 mutations per year. That might not sound like a lot, but it is important to remember that influenza viruses replicate very quickly, producing billions of copies in a single infected host. Each of these copies is an opportunity for a new mutation to arise. Most of these mutations will be neutral or deleterious, but occasionally, a mutation will confer a fitness advantage - like the ability to evade the immune system or bind to a new host receptor. Over time, these beneficial mutations can accumulate, potentially leading to the emergence of a pandemic strain.

#### Reassortment Evidence (Initial Score: 4.0)

H5N1 has demonstrated a high propensity for reassortment, both among avian influenza strains and with human seasonal influenza viruses. Numerous studies have documented reassortant H5N1 strains in birds,^12^ and *in vitro* experiments have shown that H5N1 can readily reassort with human H3N2 viruses.^13^ The ability to acquire novel gene segments through reassortment is a key driver of influenza pandemics, making this a high-risk trait for H5N1.

Let us revisit our cookbook analogy to understand the implications of reassortment. Imagine that an H5N1 “cookbook” has a page with instructions for making a dish that humans have never tasted before (i.e., a novel hemagglutinin (HA) gene segment). If this page gets mixed into a human influenza “cookbook,” the result could be a new recipe that the human immune system does not recognize. This is essentially what happened in the 1957 and 1968 pandemics, where reassortment events introduced avian HA and PB1 gene segments into human-adapted influenza viruses.^1,2^

#### Incubation Period (Initial Score: 4.0)

The incubation period for H5N1 in humans is estimated to be around 2 to 5 days, based on limited data from confirmed cases.^14^ This is shorter than the incubation period for some other zoonotic viruses like MERS-CoV and Nipah virus, but longer than that of human seasonal influenza viruses.^15–17^ A longer incubation period can make it more difficult to detect and control outbreaks, as infected individuals may spread the virus before developing symptoms. However, the relatively small number of human cases to date means that there is still considerable uncertainty around this trait. We assigned an initial score of 4.0, indicating a moderate-to-high level of risk.

The incubation period is an important factor in the controllability of a pandemic virus. A virus with a very short incubation period, like norovirus (which can be as short as 12 hours),^18^ can spread explosively, as infected individuals can transmit the virus before they even realize they are sick. On the other hand, a virus with a very long incubation period, like HIV (which can be years),^19^ can spread silently for an extended period before being detected. H5N1’s estimated incubation period of 2-5 days falls somewhere in the middle. It is long enough that infected individuals could potentially travel and expose others before developing symptoms, but short enough that outbreaks could still be identified and controlled with rapid response measures.

#### Transmission Mode: Zoonotic Transmission (Initial Score: 4.0)

H5N1 has a well-documented ability to cross the species barrier from birds to humans. As of December 2024, the WHO has reported a total of 954 confirmed human cases of H5N1,^5^ all of which are believed to have originated from exposure to infected poultry. The virus has also been detected in a range of other mammalian hosts, including pigs, cats, and most recently, cattle.^20^ However, after expert discussion, the initial score of 4.0 was revised to 3.5 to better reflect the sporadic nature of these transmissions and the observation that despite widespread avian outbreaks, human cases remain relatively rare. This pattern suggests that while cross-species transmission is possible, there are still significant barriers to efficient zoonotic spread.

The zoonotic transmission of avian influenza viruses to humans is a major concern because it is essentially a numbers game. The more times a virus jumps from birds to humans, the more chances it has to acquire mutations that allow it to replicate effectively in human cells and spread between human hosts. Each zoonotic spillover event is like a roll of the evolutionary dice. Most of the time, the virus will hit a dead end, unable to sustain transmission in the new host. But if the right combination of mutations comes up, the result could be a pandemic strain.

#### Demographic Impact (Initial Score: 3.0)

The demographic profile of human H5N1 cases shows a disproportionate impact on children and young adults, with a median age of 18 years.^21^ This is a concerning pattern, as high morbidity and mortality in younger age groups can have severe societal and economic consequences. However, the overall demographic impact of H5N1 to date has been limited by the small total number of cases. If the virus were to acquire the ability to spread efficiently among humans, the demographic impact would likely be much higher. We assigned an initial score of 3.0 for this trait, reflecting a moderate level of risk based on the current epidemiological data.

The demographic impact of a pandemic can be just as important as its overall severity in terms of societal disruption. The 1918 influenza pandemic, for example, had a peculiar mortality pattern, with a peak in young adults (20-40 years old) in addition to the usual peaks in young children and the elderly.^4^ This led to significant economic and social disruption, as the young adult population is typically the most productive segment of the workforce. The fact that H5N1 seems to disproportionately affect children and young adults raises similar concerns, although the total number of cases so far is too small to draw definitive conclusions.

#### Aerosol Transmission (Initial Score: 2.5)

The ability of H5N1 to transmit via aerosols or respiratory droplets represents a critical factor in its pandemic potential. Current evidence suggests that while H5N1 can occasionally spread through close-contact respiratory routes in experimental settings, efficient aerosol transmission between humans has not been demonstrated.^21^ Laboratory studies using ferrets, the gold-standard animal model for human influenza transmission, have shown that specific adaptive mutations are required for H5N1 to achieve efficient respiratory transmission. These mutations primarily affect the viral hemagglutinin protein, enabling better binding to human-type receptors in the upper respiratory tract.^22,23^

The score of 2.5 reflects the virus’s current limited capacity for aerosol spread while acknowledging its demonstrated potential to acquire this ability through specific genetic changes. This assessment is supported by epidemiological data showing that most human H5N1 infections result from direct contact with infected birds rather than respiratory transmission between people.^22,23^ However, the identification of mutations that can enable respiratory droplet transmission in experimental settings suggests that this barrier to pandemic spread could potentially be overcome through continued evolution. This possibility is particularly concerning given H5N1’s high mutation rate and frequent opportunities for adaptation during zoonotic spillover events.

#### Human-to-Human Spread (Initial Score: 2.0)

To date, H5N1 has demonstrated limited transmissibility between humans. The vast majority of human cases have been linked to direct exposure to infected birds, with only rare instances of limited human-to-human transmission.^21^ However, the virus has shown the ability to bind to human-type receptors in the upper respiratory tract,^24^ and ferret studies have demonstrated that a small number of mutations can enable respiratory droplet transmission.^22^ While the current risk of sustained human transmission is low, the potential for the virus to acquire this ability through mutation or reassortment cannot be discounted.

Let us unpack what “limited human-to-human transmission” means. In the context of H5N1, this typically refers to cases where a person who had close, prolonged contact with an infected individual - usually a family member - also became infected. These secondary cases are often dead ends, meaning that the chain of transmission ends with them. For a virus to have pandemic potential, it needs to be able to sustain transmission across multiple “generations” of cases, spreading efficiently from person to person. H5N1 has not demonstrated this ability yet, but the fact that it can infect humans at all and has occasionally spread between them keeps the door open for further adaptation.^21^

#### Asymptomatic Infection (Initial Score: 1.5)

The extent of asymptomatic H5N1 infection in humans is not well characterized, but several studies suggest that it may be more common than initially thought. Seroprevalence surveys in poultry workers and other high-risk groups have found evidence of H5N1 antibodies in individuals with no history of severe illness.^11^ However, the interpretation of these findings is complicated by the possibility of cross-reactivity with other influenza subtypes. Given these substantial uncertainties and the relative rarity of documented asymptomatic cases, we assigned a conservative initial score of 1.5.

Asymptomatic infections are a double-edged sword when it comes to pandemic risk. On one hand, they can facilitate the silent spread of a virus, as infected individuals may continue to go about their daily lives, unknowingly exposing others. On the other hand, asymptomatic cases are by definition milder, and a virus that causes a high proportion of asymptomatic infections may be less likely to overwhelm healthcare systems or cause significant morbidity and mortality. For H5N1, while the possibility of asymptomatic infections cannot be ruled out, current evidence suggests they are relatively rare compared to symptomatic cases. The key question remains whether the limited seroprevalence data truly reflect asymptomatic infections or merely cross-reactivity with other influenza strains. More targeted studies with improved specificity and longitudinal follow-up are needed to resolve this uncertainty.

These initial scores were based on a comprehensive review of the available literature, but we recognized that expert input was necessary to refine the assessments and ensure that the latest scientific understanding was reflected. The final scores incorporate the feedback and insights gained through the structured expert elicitation process, as described in the **Methods** section.

### 3. Extended Results

#### 3.1. Initial and Refined H5N1 Scores

**Supplementary Table S1** details the initial literature-based H5N1 scores (pre-expert refinement) and the final, consensus-based values. Notably, Transmission Mode dropped from 4.0 to 3.5, while other categories remained the same or only slightly changed.

**Supplementary Table S1.**
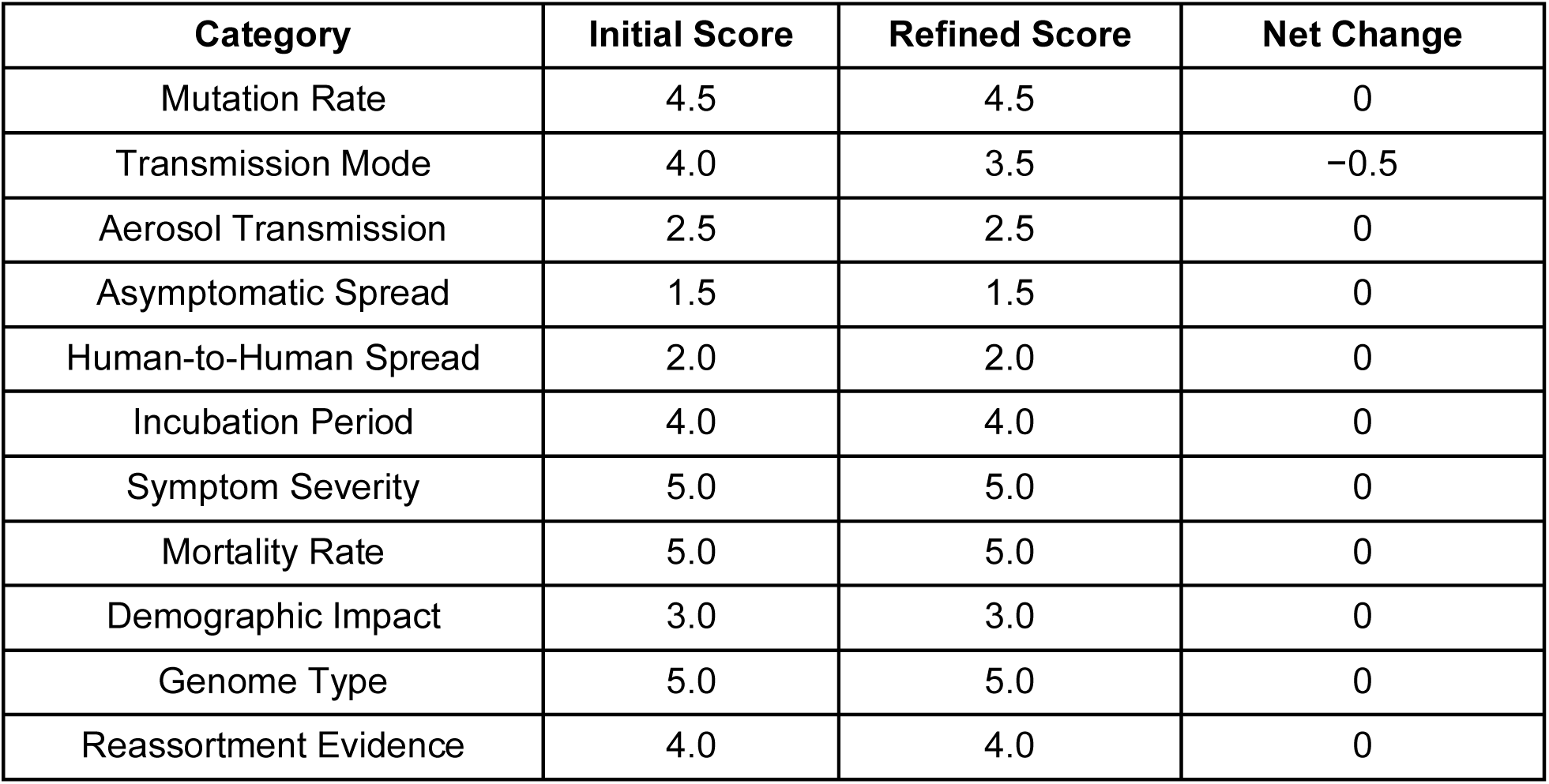
Initial vs. Refined H5N1 ViTAP Scores.

##### Supplementary Figures

**Supplementary Figure 1.**
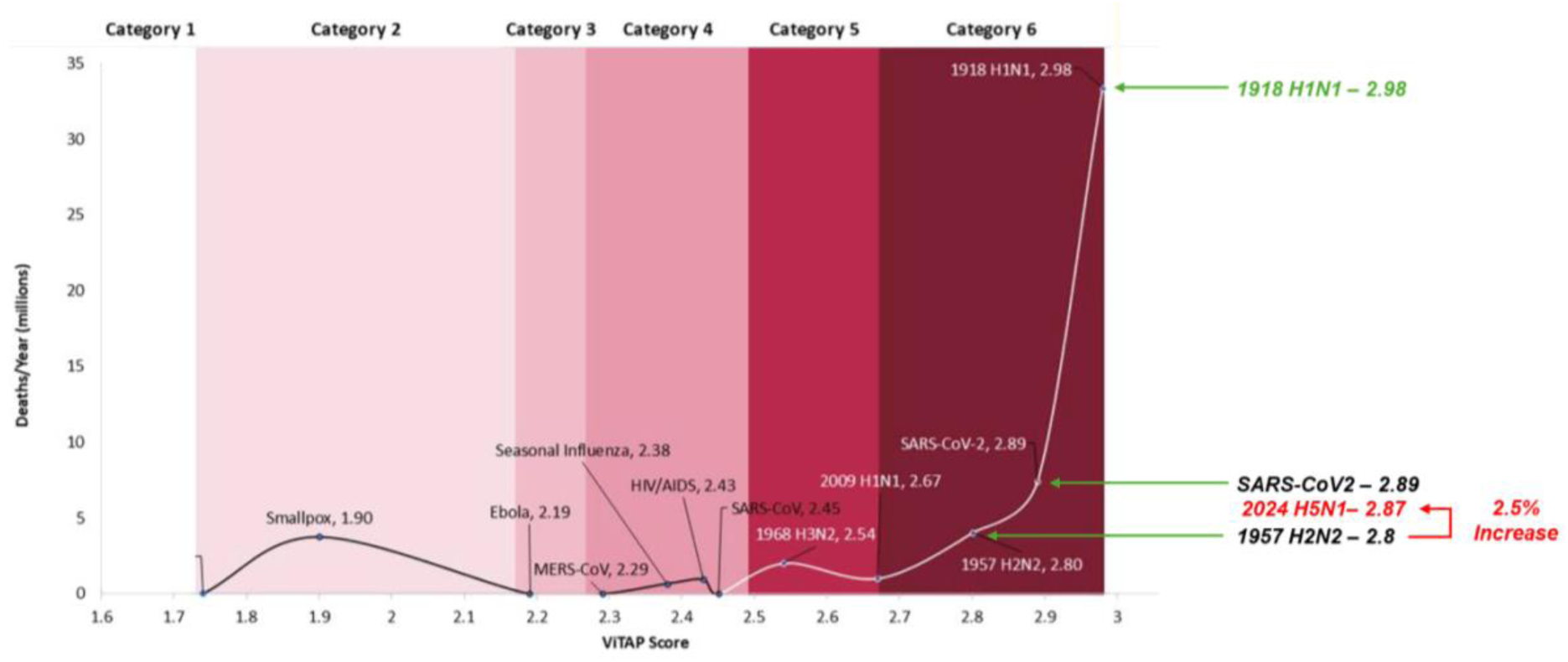

