## Supplementary material for "Assessing the Pandemic Potential of Emerging Avian Influenza A(H5N1) in the United States Using the Viral Trait Assessment for Pandemics (ViTAP) Model": SI

**3. Extended Results**

**3.1. Initial and Refined H5N1 Scores**

**Supplementary Table S1** details the initial literature-based H5N1 scores (pre-expert refinement) and the final, consensus-based values. Notably, Transmission Mode dropped from 4.0 to 3.5, while other categories remained the same or only slightly changed.

**Supplementary Table S1.** *Initial vs. Refined H5N1 ViTAP Scores*

| **Category** | **Initial Score** | **Refined Score** | **Net Change** |
| --- | --- | --- | --- |
| Mutation Rate | 4.5 | 4.5 | 0 |
| Transmission Mode | 4.0 | 3.5 | −0.5 |
| Aerosol Transmission | 2.5 | 2.5 | 0 |
| Asymptomatic Spread | 1.5 | 1.5 | 0 |
| Human-to-Human Spread | 2.0 | 2.0 | 0 |
| Incubation Period | 4.0 | 4.0 | 0 |
| Symptom Severity | 5.0 | 5.0 | 0 |
| Mortality Rate | 5.0 | 5.0 | 0 |
| Demographic Impact | 3.0 | 3.0 | 0 |
| Genome Type | 5.0 | 5.0 | 0 |
| Reassortment Evidence | 4.0 | 4.0 | 0 |

**Supplementary Figures**

**Supplementary Figure 1**

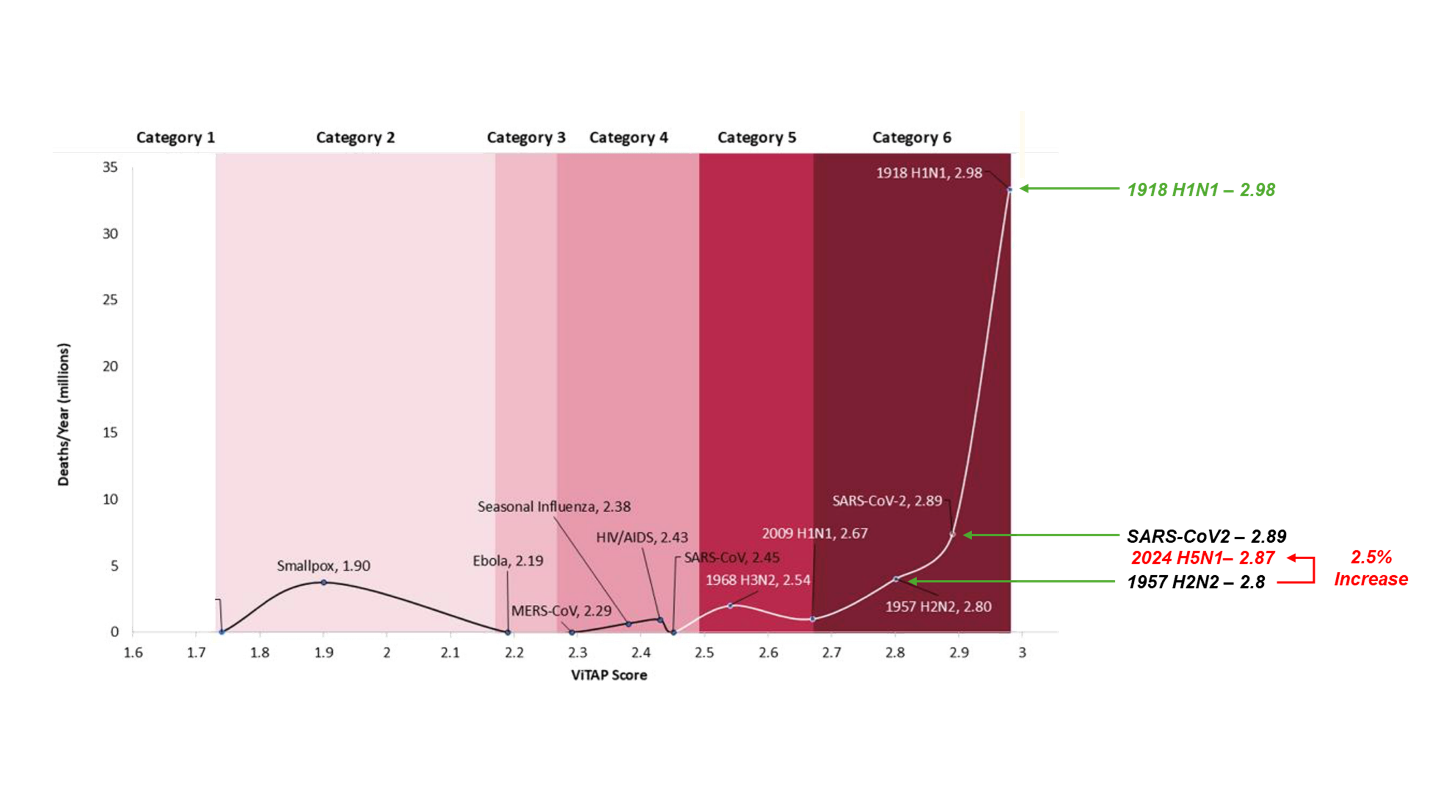
